## Supplementary material 1 for "POST-COVID ORTHOPAEDIC ELECTIVE RESOURCE PLANNING USING SIMULATION MODELLING"

**STRESS-DES: Strengthening the Reporting of Empirical Simulation Studies**

A standardised checklist to improve the reporting of discrete-event simulation models

Monks, T., Currie, C. S., Onggo, B. S., Robinson, S., Kunc, M., & Taylor, S. J. (2019). Strengthening the reporting of empirical simulation studies: Introducing the STRESS guidelines. Journal of Simulation, 13(1), 55-67. <https://doi.org/10.1080/17477778.2018.1442155>

**1.0 Objectives**

**1.1 Purpose of the model**

The HEP orthopaedic capacity planning model provides a tool for healthcare planners, managers, and clinicians to support capacity and efficiency decisions for orthopaedic services. It includes planning for two classes of surgery:

- Primary
- Revision

With five surgical types:

- Primary total hip replacement
- Primary total knee replacement
- Primary unicompartmental knee replacement
- Revision total hip replacement
- Revision total knee replacement

**1.2 Model outputs**

For each scenario configuration, the model outputs:

(i) Bed utilisation (mean/CI per day/weekday)

(ii) Total mean surgical throughput (by surgical type).

(iii) 'Lost theatre slots' which represent a mismatch between demand (patients scheduled for surgery), and capacity (beds available)

**1.3 Experimental aims**

A set of 72 indicative experimental parameter configurations were investigated:

Bed numbers between 30 and 70 beds (in increments of 5 beds; a total of 9 bed parameters) were investigated with each of two theatre schedules. The Baseline theatre schedule (4 theatres, 3 sessions per day: (i) revision *or* two primaries; (ii) revision *or* two primaries; (iii) one primary) with 5-day working. The Baseline + weekend schedule uses the same daily theatre allocations, and theatre numbers, with 7-day working. For each of these parameter changes, four scenarios were run for lengths-of-stay (baseline, 25% baseline) and for proportion delayed (baseline, 25% baseline). This totals 72 combinations. Reducing lengths-of-stay and proportion with a discharge delay by 75% brings them in line with national benchmarks. Other scenarios for increasing theatre capacity and changing theatre activity can additionally be investigated in the model.

**2.0 Model logic**

**2.1 Base model overview diagram**

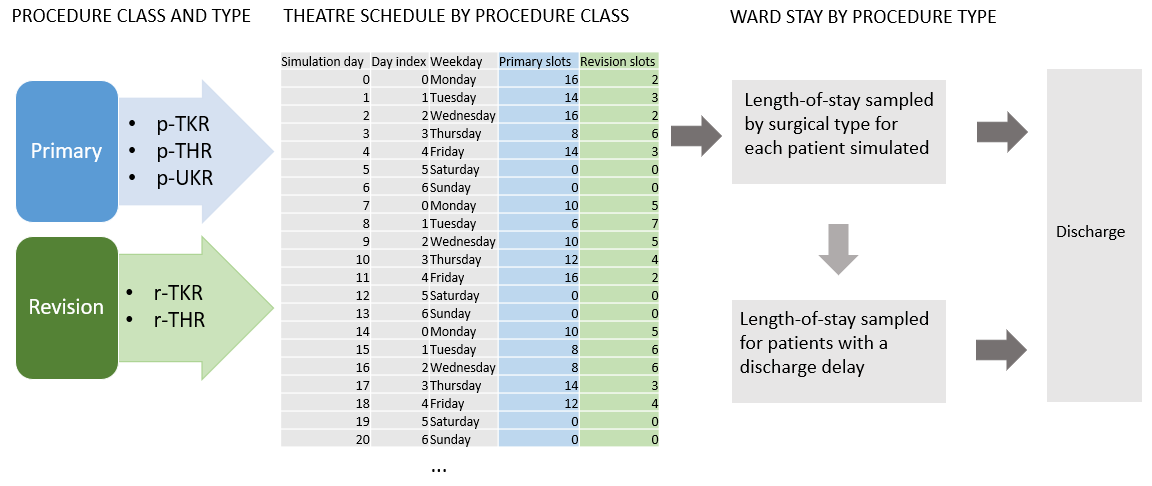

**2.2 Base model logic**

Patients arrive into the model deterministically using a pre-defined theatre schedule which allocates surgical classes (primary, revision) across a number of sessions per weekday, and a number of theatres per weekday. A surgical type is allocated by probability. Each surgical type samples a length-of-stay from a lognormal distribution fitted to historical data. A proportion of patients will have a delay to their discharge, for example waiting for out-of-hospital care. These patients have a length-of-stay sample from a lognormal distribution. If there is no bed available for a patient scheduled for surgery, this patient represents a 'lost slot'.

**2.3 Scenario logic**

- Patients arrive into the model according to a pre-defined theatre schedule. The schedule is an experimental parameter. Per weekday, the number of surgery sessions, the allocation of surgical classes per session, and the number of theatres available can be defined.
- Within the model which is made available in streamlit.io, the simulation parameters which can be changed are: number of beds; mean lengths-of-stay per surgical procedure; mean length-of-stay of patients whose discharge is delayed; proportion of patients whose discharge is delayed. Each simulation scenario can be run with the baseline theatre schedule alone, or additionally with a user-defined schedule.
- Within the model notebook, additional parameters can be changed. These include: stdev of all lengths of stay; proportions of surgical types; length-of-stay distributions.

**2.4 Components**

- A theatre schedule defines deterministic arrivals by patient class
- Patients renege after 0.5-1 day (uniform) if no bed is available (‘lost slots’)
- Each class has attributes associated with surgical procedure, or delayed discharge
- A single service node
- A single resource: beds
- FIFO queue discipline

**3.0 Data**

Routinely collected data from North Bristol NHS Trust was used to identify patients receiving elective joint arthroplasty between January 2016 and December 2019.

The Trust’s electronic health records (EHR) were used to identify elective joint arthroplasty using a combination of OPCS4 procedure and surgical site codes. Five core elective orthopedic surgical procedure types were identified and verified. A small number of short day-case ‘hip resurfacing’ surgeries [n=52] were removed from the dataset. The five remaining surgical types were classified into two classes: (I) Primary (primary hip replacement [p-THR n=3057; 51%], primary knee replacement [p-TKR; n=2302; 38%], uni-compartmental knee replacement [p-UKR; n=679; 11%]); (II) Revision (revision hip replacement [r-THR; n=482; 55%], revision knee replacement [r-TKR; n=392; 45%]). Most patients did not remain in hospital once they were medically fit for discharge, however a proportion of patients in the EHR had a recorded medically fit for discharge date which preceded their actual discharge date (n=529; 7.6%).

A statistical probability distribution was fitted to each type of surgical procedure using the EHR data. The fitted length-of-stay parameters [procedure, mean days (u), standard deviation days (sd)] are: p-THR, u=4.4, sd=2.9 days; p-TKR u=4.7 days, sd=2.8; p-UKR u=2.9 days, sd=2.1; r-THR, u=6.9, sd=7.0; r-TKR, u=7.2, sd=7.6; delayed discharge, u=16.5; sd=15.1).

**4.0 Experimentation**

**4.1 Initialisation**

The model uses a warm-up period of 35 days based on visual inspection of outputs. The model is non-terminating.

**4.2 Run Length**

The model runs for 70 days. This can be changed by the user. Multiple independent replications are undertaken to control confidence intervals. The number of replications is determined using MORE plots, and is set to 50 by default.

Nelson, B. L. (2008) The MORE plot: Displaying measures of risk & error from simulation output. In *2008 Winter Simulation Conference* (pp. 413-416). IEEE.

**5.0 Implementation**

**5.1 Software of programming language**

The simulation was developed using Python 3.8. A conda software environment is used to manage versions on a local machine. Software versions are:

- jupyterlab=3.4.6

- jupyterlab-spellchecker=0.7.2

- matplotlib=3.5.3

- numpy=1.23.2

- pandas=1.4.4

- pip=22.2.2

- python=3.8.12

- scipy=1.9.1

- simpy=4.0.1

- streamlit==1.17.0

**5.2 Random sampling**

All sampling uses `numpy.random.Generator`. Common random number streams are used in the model, created through seed vectors (one seed for each activity in each replication).

**5.3 Model execution**

Simpy implements a process-based simulation worldview.

**5.4 System specification**

The model was run on Intel i7-12700H CPU with 32GB RAM running Pop!_OS 22.04 Linux.

**6.0 Code access**

Code, input parameters and output data used for results in this paper are available on GitHub: [AliHarp/HEP: Model of orthopaedic ward (github.com)](https://github.com/AliHarp/HEP)

They are permanently archived using Zenodo ([HEP | Zenodo](https://zenodo.org/record/7951080)).

All are licensed using MIT permissive license [MIT License | Choose a License](https://choosealicense.com/licenses/mit/).
