## Supplementary material 2 for "POST-COVID ORTHOPAEDIC ELECTIVE RESOURCE PLANNING USING SIMULATION MODELLING"

**Table S1: OPCS-4 codes used to identify hip and knee arthroplasty surgery**

| **PRIMARY TOTAL HIP REPLACEMENT** | |  |
| --- | --- | --- |
| 4 Character Code | Description |  |
| W37.1 | Primary total prosthetic replacement of hip joint using cement |  |
| W37.8 | Other specified total prosthetic replacement of hip joint using cement |  |
| W37.9 | Unspecified total prosthetic replacement of hip joint using cement |  |
| W38.1 | Primary total prosthetic replacement of hip joint not using cement |  |
| W38.8 | Other specified total prosthetic replacement of hip joint not using cement |  |
| W38.9 | Unspecified total prosthetic replacement of hip joint not using cement |  |
| W39.1 | Primary total prosthetic replacement of hip joint NEC |  |
| W39.8 | Other specified other total prosthetic replacement of hip joint |  |
| W39.9 | Unspecified other total prosthetic replacement of hip joint |  |
| W43.1 | Primary total prosthetic replacement of other joint using cement NEC |  |
| W43.8 | Other specified total prosthetic replacement of other joint using cement NEC |  |
| W43.9 | Unspecified total prosthetic replacement of other joint using cement NEC |  |
| W44.1 | Primary total prosthetic replacement of other joint not using cement NEC |  |
| W44.8 | Other specified total prosthetic replacement of other joint not using cement NEC |  |
| W44.9 | Unspecified total prosthetic replacement of other joint not using cement NEC |  |
| W45.1 | Other primary total prosthetic replacement of other joint NEC |  |
| W45.8 | Other specified total prosthetic replacement of other joint NEC |  |
| W45.9 | Unspecified total prosthetic replacement of other joint NEC |  |
| W52.1 | Primary prosthetic replacement of articulation of bone using cement NEC |  |
| W52.8 | Other specified prosthetic replacement of articulation of bone using cement NEC |  |
| W52.9 | Unspecified prosthetic replacement of articulation of bone using cement NEC |  |
| W53.1 | Primary prosthetic replacement of articulation of bone not using cement NEC |  |
| W53.8 | Other specified prosthetic replacement of articulation of bone not using cement NEC |  |
| W53.9 | Unspecified prosthetic replacement of articulation of bone not using cement NEC |  |
| W54.1 | Primary prosthetic replacement of articulation of bone NEC |  |
| W54.8 | Other specified prosthetic replacement of articulation of bone NEC |  |
| W54.9 | Unspecified prosthetic replacement of articulation of bone NEC |  |
| W93.1 | Primary hybrid prosthetic replacement of hip joint using cemented acetabular component |  |
| W93.8 | Other specified hybrid prosthetic replacement of hip joint using cemented acetabular component |  |
| W93.9 | Unspecified hybrid prosthetic replacement of hip joint using cemented acetabular component |  |
| W94.1 | Primary hybrid prosthetic replacement of hip joint using cemented femoral component |  |
| W94.8 | Other specified hybrid prosthetic replacement of hip joint using cemented femoral component |  |
| W94.9 | Unspecified hybrid prosthetic replacement of hip joint using cemented femoral component |  |
| W95.1 | Primary hybrid prosthetic replacement of hip joint using cement NEC |  |
| W95.8 | Other specified hybrid prosthetic replacement of hip joint using cement |  |
| W95.9 | Unspecified hybrid prosthetic replacement of hip joint using cement |  |
| **RESURFACING / RECONSTRUCTION** | |  |
| 4 Character Code | Description |  |
| W58.1 | Primary resurfacing arthroplasty of joint |  |
| W58.8 | Other specified reconstruction of joint |  |
| W58.9 | Unspecified other reconstruction of joint |  |
| **LOCATION OF SURGERY** | |  |
| 4 Character Code | Description |  |
| Z75.6 | Acetabulum |  |
| Z76.1 | Head of femur |  |
| Z84.3 | Hip joint |  |
| **REVISION HIP REPLACEMENT** | |  |
| 4 Character Code | Description |  |
| W37.0 | Conversion from previous cemented total prosthetic replacement of hip joint |  |
| W37.2 | Conversion to total prosthetic replacement of hip joint using cement |  |
| W37.3 | Revision of total prosthetic replacement of hip joint using cement |  |
| W37.4 | Revision of one component of total prosthetic replacement of hip joint using cement |  |
| W38.0 | Conversion from previous uncemented total prosthetic replacement of hip joint |  |
| W38.2 | Conversion to total prosthetic replacement of hip joint not using cement |  |
| W38.3 | Revision of total prosthetic replacement of hip joint not using cement |  |
| W38.4 | Revision of one component of total prosthetic replacement of hip joint not using cement |  |
| W39.0 | Conversion from previous total prosthetic replacement of hip joint NEC |  |
| W39.2 | Conversion to total prosthetic replacement of hip joint NEC |  |
| W39.3 | Revision of total prosthetic replacement of hip joint NEC |  |
| W39.5 | Revision of one component of total prosthetic replacement of hip joint NEC |  |
| W43.0 | Conversion from previous cemented total prosthetic replacement of joint NEC | + Site code |
| W43.2 | Conversion to total prosthetic replacement of joint using cement NEC | + Site code |
| W43.3 | Revision of total prosthetic replacement of joint using cement NEC | + Site code |
| W43.4 | Revision of one component of total prosthetic replacement of joint using cement NEC | + Site code |
| W44.0 | Conversion from previous uncemented total prosthetic replacement of joint NEC | + Site code |
| W44.2 | Conversion to total prosthetic replacement of joint not using cement NEC | + Site code |
| W44.3 | Revision of total prosthetic replacement of joint not using cement NEC | + Site code |
| W44.4 | Revision of one component of total prosthetic replacement of joint not using cement NEC | + Site code |
| W45.0 | Conversion from previous total prosthetic replacement of joint NEC | + Site code |
| W45.2 | Conversion to total prosthetic replacement of joint NEC | + Site code |
| W45.3 | Revision of total prosthetic replacement of joint NEC | + Site code |
| W45.5 | Revision of one component of total prosthetic replacement of joint NEC | + Site code |
| W46.2 | Conversion to prosthetic replacement of head of femur using cement |  |
| W46.3 | Revision of prosthetic replacement of head of femur using cement |  |
| W47.2 | Conversion to prosthetic replacement of head of femur not using cement |  |
| W47.3 | Revision of prosthetic replacement of head of femur not using cement |  |
| W48.2 | Conversion to prosthetic replacement of head of femur NEC |  |
| W48.3 | Revision of prosthetic replacement of head of femur NEC |  |
| W52.0 | Conversion from previous cemented prosthetic replacement of articulation of bone NEC | + Site code |
| W52.2 | Conversion to prosthetic replacement of articulation of bone using cement NEC | + Site code |
| W52.3 | Revision of prosthetic replacement of articulation of bone using cement NEC | + Site code |
| W53.0 | Conversion from previous uncemented prosthetic replacement of articulation of bone NEC | + Site code |
| W53.2 | Conversion to prosthetic replacement of articulation of bone not using cement NEC | + Site code |
| W53.3 | Revision of prosthetic replacement of articulation of bone not using cement NEC | + Site code |
| W54.0 | Conversion from previous prosthetic replacement of articulation of bone NEC | + Site code |
| W54.2 | Conversion to prosthetic replacement of articulation of bone NEC | + Site code |
| W54.3 | Revision of prosthetic replacement of articulation of bone NEC | + Site code |
| W57.4 | Conversion to excision arthroplasty of joint | + Site code |
| W58.0 | Conversion from previous resurfacing arthroplasty of joint | + Site code |
| W58.2 | Revision of resurfacing arthroplasty of joint | + Site code |
| W93.0 | Conversion from previous hybrid prosthetic replacement of hip joint using cemented acetabular component |  |
| W93.2 | Conversion to hybrid prosthetic replacement of hip joint using cemented acetabular component |  |
| W93.3 | Revision of hybrid prosthetic replacement of hip joint using cemented acetabular component |  |
| W94.0 | Conversion from previous hybrid prosthetic replacement of hip joint using cemented femoral component |  |
| W94.2 | Conversion to hybrid prosthetic replacement of hip joint using cemented femoral component |  |
| W94.3 | Revision of hybrid prosthetic replacement of hip joint using cemented femoral component |  |
| W95.0 | Conversion from previous hybrid prosthetic replacement of hip joint using cement NEC |  |
| W95.2 | Conversion to hybrid prosthetic replacement of hip joint using cement NEC |  |
| W95.3 | Revision of hybrid prosthetic replacement of hip joint using cement NEC |  |

| **PRIMARY TOTAL KNEE REPLACEMENT** |  |  |
| --- | --- | --- |
| 4 Character Code | Description |  |
| W40.1 | Primary total prosthetic replacement of knee joint using cement |  |
| W40.8 | Other specified total prosthetic replacement of knee joint using cement |  |
| W40.9 | Unspecified total prosthetic replacement of knee joint using cement |  |
| W41.1 | Primary total prosthetic replacement of knee joint not using cement |  |
| W41.8 | Other specified total prosthetic replacement of knee joint not using cement |  |
| W41.9 | Unspecified total prosthetic replacement of knee joint not using cement |  |
| W42.1 | Primary total prosthetic replacement of knee joint NEC |  |
| W42.8 | Other specified other total prosthetic replacement of knee joint |  |
| W42.9 | Unspecified other total prosthetic replacement of knee joint |  |
| O18.1 | Primary hybrid prosthetic replacement of knee joint using cement |  |
| O18.8 | Other specified hybrid prosthetic replacement of knee joint using cement |  |
| O18.9 | Unspecified hybrid prosthetic replacement of knee joint using cement |  |
| **PRIMARY UNICONDYLAR / UNICOMPARTMENTAL KNEE OPERATIONS** | |  |
| 4 Character Code | Description |  |
| W52.1 | Primary prosthetic replacement of articulation of bone using cement NEC | Require combination with site + combination codes to ID |
| W52.8 | Other specified prosthetic replacement of articulation of other bone using cement | Require combination with site + combination codes to ID |
| W52.9 | Unspecified prosthetic replacement of articulation of other bone using cement | Require combination with site + combination codes to ID |
| W53.1 | Primary prosthetic replacement of articulation of bone not using cement NEC | Require combination with site + combination codes to ID |
| W53.9 | Unspecified prosthetic replacement of articulation of other bone not using cement | Require combination with site + combination codes to ID |
| W54.0 | Conversion from previous prosthetic replacement of articulation of bone NEC | Require combination with site + combination codes to ID |
| W54.1 | Primary prosthetic replacement of articulation of bone NEC | Require combination with site + combination codes to ID |
| W54.8 | Other specified other prosthetic replacement of articulation of other bone | Require combination with site + combination codes to ID |
| W54.9 | Unspecified other prosthetic replacement of articulation of other bone | Require combination with site + combination codes to ID |
| W58.1 | Primary resurfacing arthroplasty of joint | Require combination with site + combination codes to ID |
| **SITE OF SURGERY** |  |  |
| Z76.5 | Lower end of femur NEC |  |
| Z77.4 | Upper end of tibia NEC |  |
| Z78.7 | Patella | Will need care in extracting as PFJ replacement will be coded with this |
| Z84.4 | Patellofemoral joint | Will need care in extracting as PFJ replacement will be coded with this |
| Z84.5 | Tibiofemoral joint |  |
| Z84.6 | Knee joint |  |
| **REVISION KNEE REPLACEMENT** |  |  |
| 4 Character Code | Description |  |
| W40.0 | Conversion from previous cemented total prosthetic replacement of knee joint |  |
| W40.2 | Conversion to total prosthetic replacement of knee joint using cement |  |
| W40.3 | Revision of total prosthetic replacement of knee joint using cement |  |
| W40.4 | Revision of one component of total prosthetic replacement of knee joint using cement |  |
| W41.0 | Conversion from previous uncemented total prosthetic replacement of knee joint |  |
| W41.2 | Conversion to total prosthetic replacement of knee joint not using cement |  |
| W41.3 | Revision of total prosthetic replacement of knee joint not using cement |  |
| W41.4 | Revision of one component of total prosthetic replacement of knee joint not using cement |  |
| W42.0 | Conversion from previous total prosthetic replacement of knee joint NEC |  |
| W42.2 | Conversion to total prosthetic replacement of knee joint NEC |  |
| W42.3 | Revision of total prosthetic replacement of knee joint NEC |  |
| W42.4 | Attention to total prosthetic replacement of knee joint NEC | Plus Y03.2 (Renewal of prosthesis in organ NOC) or Y03.7 (Removal of prosthesis from organ NOC) |
| W42.5 | Revision of one component of total prosthetic replacement of knee joint NEC |  |
| W42.6 | Arthrolysis of total prosthetic replacement of knee joint |  |
| W58.0 | Conversion from previous resurfacing arthroplasty of joint |  |
| W58.2 | Revision of resurfacing arthroplasty of joint | Require combination with site + combination codes to ID |
| O18.0 | Conversion from previous hybrid prosthetic replacement of knee joint using cement |  |
| O18.2 | Conversion to hybrid prosthetic replacement of knee joint using cement |  |
| O18.3 | Revision of hybrid prosthetic replacement of knee joint using cement |  |
| O18.4 | Attention to hybrid prosthetic replacement of knee joint using cement |  |
| W52.0 | Conversion from previous cemented prosthetic replacement of articulation of bone NEC | Require combination with site + combination codes to ID |
| W52.2 | Conversion to prosthetic replacement of articulation of bone using cement NEC | Require combination with site + combination codes to ID |
| W52.3 | Revision of prosthetic replacement of articulation of bone using cement NEC | Require combination with site + combination codes to ID |
| W53.0 | Conversion from previous uncemented prosthetic replacement of articulation of bone NEC | Require combination with site + combination codes to ID |
| W53.2 | Conversion to prosthetic replacement of articulation of bone not using cement NEC | Require combination with site + combination codes to ID |
| W53.3 | Revision of prosthetic replacement of articulation of bone not using cement NEC | Require combination with site + combination codes to ID |
| W54.0 | Conversion from previous prosthetic replacement of articulation of bone NEC | Require combination with site + combination codes to ID |
| W54.2 | Conversion to prosthetic replacement of articulation of bone NEC | Require combination with site + combination codes to ID |
| W54.3 | Revision of prosthetic replacement of articulation of bone NEC | Require combination with site + combination codes to ID |
| W54.4 | Attention to prosthetic replacement of articulation of bone NEC | Require combination with site + combination codes to ID |
| W55.3 | Conversion to prosthetic interposition arthroplasty of joint | Require combination with site + combination codes to ID |
| W56.4 | Conversion to interposition arthroplasty of joint NEC | Require combination with site + combination codes to ID |
| W57.4 | Conversion to excision arthroplasty of joint | Require combination with site + combination codes to ID |
| W60.3 | Conversion to arthrodesis and extra-articular bone graft NEC | Require combination with site + combination codes to ID |
| W61.3 | Conversion to arthrodesis and articular bone graft NEC | Require combination with site + combination codes to ID |
| W64.1 | Conversion to arthrodesis and internal fixation NEC | Require combination with site + combination codes to ID |
| W64.2 | Conversion to arthrodesis and external fixation NEC | Require combination with site + combination codes to ID |
